# Efficient positioning of QTL and Secondary Limit thresholds in a clinical trial risk-based monitoring

**DOI:** 10.1101/2024.06.07.24308630

**Authors:** Vladimir Shnaydman

## Abstract

In the high-stakes world of clinical trials, where a company’s multimillion-dollar drug development investment is at risk, the increasing complexity of these trials only compounds the challenges. Therefore, the development of a robust risk mitigation strategy, as a crucial component of comprehensive risk planning, is not just important but essential for effective drug development, particularly in the RBQM ecosystem. This emphasis on the urgency and significance of risk mitigation strategy can help the audience understand the gravity of the topic.

The paper introduces a novel framework for deriving an efficient risk mitigation strategy at the planning stage of a clinical trial and establishing operational rules (thresholds). This approach combines optimization and simulation models, offering a fresh perspective on risk management in clinical trials. The optimization model aims to derive an efficient contingency budget and allocate limited mitigation resources across mitigated risks. The simulation model aims to efficiently position the QTL/KRI and Secondary Limit thresholds for each risk to be aligned with risk assessment and contingency resources.

A compelling case study vividly illustrates the practical application and effectiveness of the proposed technique. This real-world example not only demonstrates the framework’s potential but also instills confidence in its successful implementation, reassuring the audience of its practicality and effectiveness.

## Introduction

A clinical trial holds many potential risks that could jeopardize a company’s multimillion-dollar drug development investment. The increasing complexity of clinical trials compounds these challenges. Therefore, comprehensive risk planning, including an efficient risk mitigation strategy, is essential for clinical trial planning and execution.

The typical risk planning process is cyclical and includes four interconnected steps: identification of potential risks, risk assessment, risk response planning, and risk monitoring.

Risk response planning aims to reduce risk. Several options could be considered: risk avoidance (an alternative approach that does not include a particular risk), risk prevention, risk transfer (e.g., insurance to cover patients’ safety), risk acceptance, and risk mitigation. The paper addresses clinical trial risk mitigation planning.

Risk mitigation planning includes three interconnected components:

1. Development of a risk mitigation strategy^1^.
2. Establishment of the operational risk mitigation rules^2^
3. Alignment between the mitigation strategy and the operational risk mitigation rules.

Four critical problems need to be addressed in the risk mitigation strategy planning.

1. Derivation of a contingency budget.
2. Efficient contingency budget allocation across risks.
3. Efficient selection of a risk mitigation option for each risk, assuming some risks may be associated with several mutually exclusive risk mitigation options (e.g., backup sites in country A or B).
4. Alignment of a contingency budget and risk tolerance

The modeling technique to address risk mitigation strategy planning was presented in [1]. The method assumes efficient risk mitigation is possible if sufficient contingency resources are allocated. However, efficient allocation of contingency resources does not guarantee flawless risk mitigation process execution. The process should be subject to operational rules that keep multiple Risk Parameters (RP) within an acceptable range to minimize risk exposure during clinical trial execution [2]. These RP are vital indicators that help us monitor and manage risks effectively. The operational rules are derivatives of the risk mitigation strategy and are defined by thresholds.

Identifying and monitoring RP is crucial for making informed decisions and implementing effective risk management strategies.

This paper defines RP as a relative risk metric [3]^3^.

There are two approaches for executing risk mitigation – reactive and proactive. The <u>reactive</u> approach assumes that risk mitigation starts when a risk becomes an issue [4, 5]. A mitigation action could be delayed by days and weeks, and the impact related to the risk eventuation will increase. The <u>proactive</u> approach assumes risk mitigation should start earlier after an RP reaches a specific “early warning” level to avoid the potential risk eventuation.

Only Quality Tolerance Limit (QTL)^4^ or Key Risk Indicator (KRI)^5^ Thresholds are needed to implement the reactive approach.^6^ For the proactive approach, in addition to QTL/KRI thresholds^7^, the Secondary Limit (SL)^8^ Threshold is necessary as an early warning system for better risk management. According to an empirical rule, the SL could be placed at 50-75% of the QTL level [5]. The rule does not reflect clinical trial specifics.

The critical issue in RBM is how to efficiently position QTL and SL thresholds and align them with risk assessment and mitigation costs.

Early warning systems, which aim to inform about risks and mitigate them before they occur, are used in many industries. For example, in seismology, they predict earthquakes [6], in hydrology, flooding [7], and the failure of IT projects [8].

Substantial research has been dedicated to establishing and positioning the QTLs in clinical trials, ranging from adapting the Statistical Process Control (SPC) methodology successfully applied in mass manufacturing [9, 10] to extrapolating historical data for similar trials [3, 4, 11, 12] and QTL benchmarking [13].

1. <u>The SPC approach</u> was developed for quality control in mass manufacturing production and based on the analysis of sampling batches [9]. In clinical trials, it was applied to COVID-19 vaccine clinical trials when only one shot was required, and the number of participants was large enough (within the 30K - 60K range) [10] to be divided into sampling batches. At the same time, the author [10] admits, *“A small trial can be difficult to monitor* (*using SPC*) *because proportions and other statistics can be imprecise due to a small sample size*.*”*
2. <u>The historical data approach</u> is based on benchmarking data from prior trials in the same therapeutic area and indication. It assumes data homogeneity [11,12]. However, even two trials in the same therapeutic area and indication may have distant risk profiles. Due to insufficient historical data, this approach may have limited value for medium and small companies. Also, new technologies, e.g., DCT/Hybrid, limit the number of comparable trials.
3. <u>QTL data benchmarking.</u> Assigning QTL thresholds based on benchmarking data could also be problematic. For Example, in [13], a range of QTLs for a predefined set of RPs was presented. The data was taken from a consortium of companies [14]. The results in [13] indicate that the QTL thresholds vary widely due to each trial’s uniqueness. The results are presented as (Average, ±Standard Deviation) can potentially include even negative values for QTL thresholds, which is hard to explain. While widely used, this approach has limitations. It may not always provide accurate or consistent results, especially in clinical trials where each trial is unique, and the risk profiles can vary significantly.
4. <u>Subjective</u> QTL threshold positioning based on previous experience.

The approaches 1-4 may have limited value because they

- Are not trial-specific.
- Are not aligned with the trial risk and risk mitigation strategy.
- Have limited value for small and medium companies with limited historical data related to comparable trials.

The paper describes a methodology based on combined optimization and simulation modeling. This approach can relax current techniques’ limitations and increase risk mitigation planning accuracy.

The paper is structured as follows. (1) Introduction; (2) Methodology; (3) Models – Risk Optimizer and Risk Simulator; (4) Results; and (5) Conclusion.

## Methodology

The methodology presented in Figure 1 is based on interconnected strategic and operational risk planning mathematical models. It includes three steps: (1) strategic risk planning (Risk Optimizer), (2) operational risk planning (Risk Simulator) – to derive RBM operational rules, and (3) alignment of strategic and operational risk planning.

**Figure 1.**
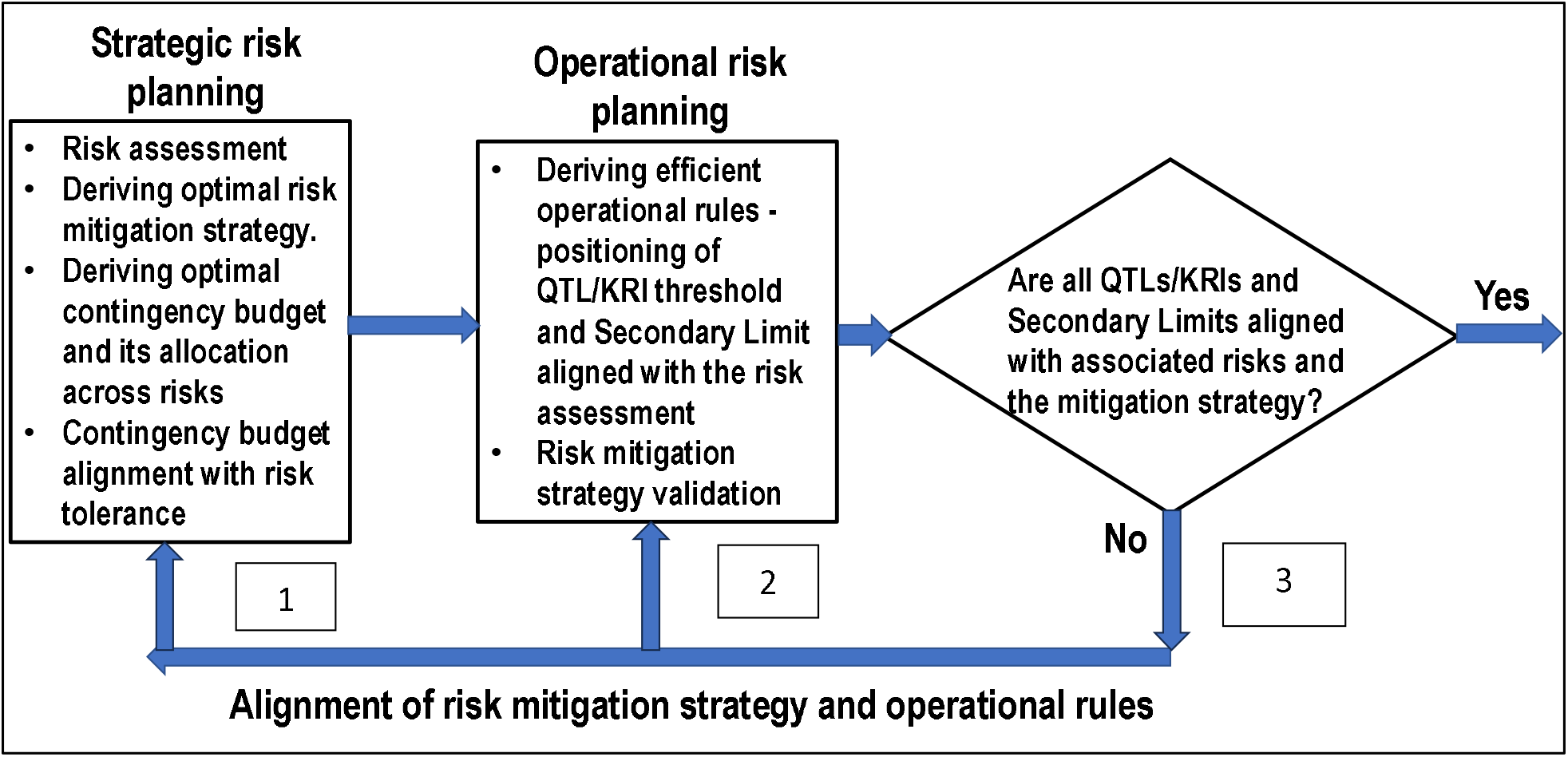
The risk mitigation planning methodology.

<u>Strategic risk planning</u> includes (1) risk assessment, (2) the optimal risk mitigation strategy planning (selection of an efficient mitigation option for each risk), (3) the optimal contingency budget derivation and its allocation across risks, and (4) contingency budget alignment with risk tolerance [1]. The outcome of the strategic risk planning stage is (a) residual risk and (b) allocated resources to mitigate each risk.

Operational risk planning goals include deriving efficient operational rules, i.e., positioning QTL and SL thresholds aligned with risk assessment and mitigation resources.

A simplified example/analogy explains the proposed methodology.

### Example

Each modern car has a fuel indicator. Suppose the fuel level in the tank reaches a low-level point where the indicator flashes. The responsible driver should stop at the nearest gas station if they have sufficient money to fill the tank to mitigate the risk of an empty tank. The risk of an empty tank in the middle of the road is considered critical and has a significant negative impact.

The driver may have several mitigation options. They are: (1) filling up the tank at the nearest gas station; (2) if fuel is expensive at the nearest gas station, they can partially fill the tank enough to get to a cheaper station; (3) passing the current gas station and fill the tank at a cheaper station, but the risk of an empty tank will increase.

If the driver faces two risks (e.g., an empty tank and a potential flat tire), they must allocate money to mitigate them - a potential tire repair/replacement and filling up the tank.

This analogy can be applied to the clinical trial risk management process.

RP is associated with the fuel amount in the tank, QTL is associated with an empty tank, and a fuel indicator is associated with SL.

Assumptions:

1. all analyzed risks are mitigated.
2. a contingency budget is limited.
3. each mitigation action requires resources.

### 1. Optimal risk mitigation strategy – Risk Optimizer [1]

An efficient risk mitigation strategy should be able to address the following questions.

a. How to derive a contingency budget that is aligned with risk?
b. How can a contingency budget be optimally allocated across risks?
c. How can the risk mitigation strategy for a clinical trial be optimized?
d. How can a contingency budget, mitigation strategy, clinical trial risk capacity, risk appetite, and risk tolerance be aligned?

## The problem statement

▪ In a clinical trial, high and medium risks should be mitigated.
▪ Each risk may have several mutually exclusive mitigation options. For example, backup clinical sites could be in countries X, Y, or both (Table 1).
▪ Each mitigation option is characterized by the likelihood score (**L**) (Table 2), impact score(**I**) (Table 3), the total score (**LxI**), mitigation costs (**M**), risk/probability (**P**) associated with the likelihood score (Table 2), and risk-adjusted mitigation costs (**MxP**).
▪ Contingency budget - **B**

### Input data

#### The optimization model

The model selects an optimal risk mitigation plan, allocating a contingency budget across risks and minimizing the Total Normalized Risk Score (TNRS – a sum of risk scores divided by a total number of mitigated risks) [1] for the entire portfolio of clinical trial risks. It also allocates limited contingency resources across risks, selecting an optimal mitigation option for each risk.

**Table 1.** Input data and modeling results (example).

| Input |  |  |  |  |  |  |  | Output |
| --- | --- | --- | --- | --- | --- | --- | --- | --- |
|  | # | Likelihood Score (**L**) | Impact Score (**I**) | Total Score (**LxI**) | Mitigation costs –(**M**) | Probability (**P**) | Risk-adjusted mitigation costs (**MxP**) | The model selected the mitigation option |
| Risk | 1 |  |  |  |  |  |  |  |
| Mitigation options | 1.1 (no mitigation) |  |  |  |  |  |  | X |
|  | 1.2 |  |  |  |  |  |  |  |
| Risk | 2 |  |  |  |  |  |  |  |
| Mitigation options | 2.1 (no mitigation) |  |  |  |  |  |  |  |
|  | 2.2 |  |  |  |  |  |  |  |
|  | 2.3 |  |  |  |  |  |  | X |

**Table 2.** The likelihood score **L**_**i**_ scaled from j = (1,5) is a surrogate for an average risk/probability **P**_**i**_ = (0,1) (Li ⟷ Pi) or the range of probabilities presented in Table 1. For example, due to assessment subjectivity, a score of five corresponds to “a very high” risk/probability of 90% or within the 85% - 95% range.

|  |  |  |  |  |  |
| --- | --- | --- | --- | --- | --- |
| Likelihood score - L | 1 | 2 | 3 | 4 | 5 |
| Qual. Rating | Very low | Low | Medium | High | Very high |
| Average, risk/probability P, % | 10 | 25 | 50 | 75 | 90 |
| Range, | 5-15 | 15-35 | 35-65 | 65-85 | 85-95 |
| risk/probability<br>($P_{\min}$, $P_{\max}$), % | | | | | |

**Table 3.** Impact score.

|  |  |  |  |  |  |
| --- | --- | --- | --- | --- | --- |
| Impact - I | 1 | 2 | 3 | 4 | 5 |
| Rating | Very low | Low | Medium | High | Very high |

### 2. RBM risk simulator

The RBM simulator mimics RP dynamics for each mitigated risk. It was developed to validate the risk mitigation strategy and establish operational rules.

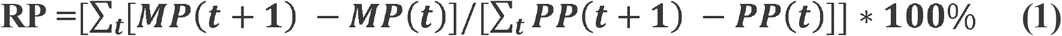

MP is the parameter’s monitored value, and PP is its planned value.

An RP dynamic is presented in (2) and described as

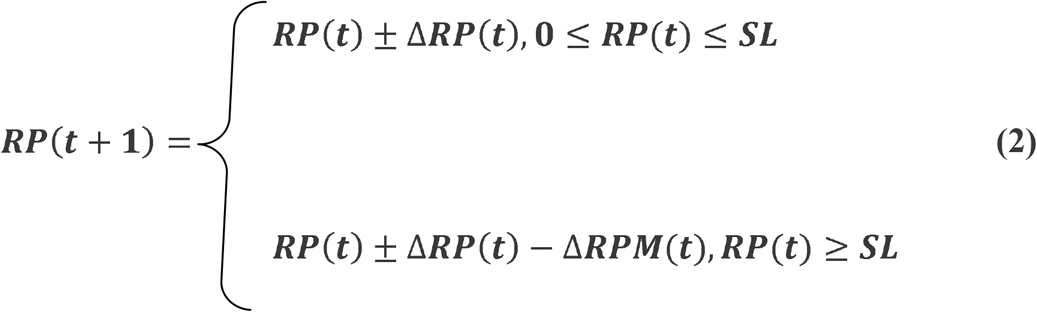

Where **RP(t)** and **RP(t+1)** are values of the RP in times **t** and (**t+1**), **Δ*RP*(*t*)** - is an increase/reduction in the value of an RP. **Δ*RP*(*t*)** is uniformly distributed within a range **(Δ*RPmin*(*t*), (Δ*RPmax*(*t*))**.

**Δ*RPM*(*t*)** is an incremental reduction of an RP in the interval **(t, t+1)** due to risk mitigation if ***RP*(*t*) *min*(*SL, QTL*). Δ*RPM*(*t*)** is also uniformly distributed within a range **(Δ*RPMmin*(*t*), (Δ*RPMmax*(*t*)**). Values of both **Δ*RP*(*t*) *and* Δ*RPM*(*t*)** are based on historical observations and expert judgment^9^. The parameter **Δ*RPM*(*t*)** It also depends on available contingency resources allocated for a mitigated risk.

An illustration of the RP dynamic is presented in Figure 2.

**Figure 2.**
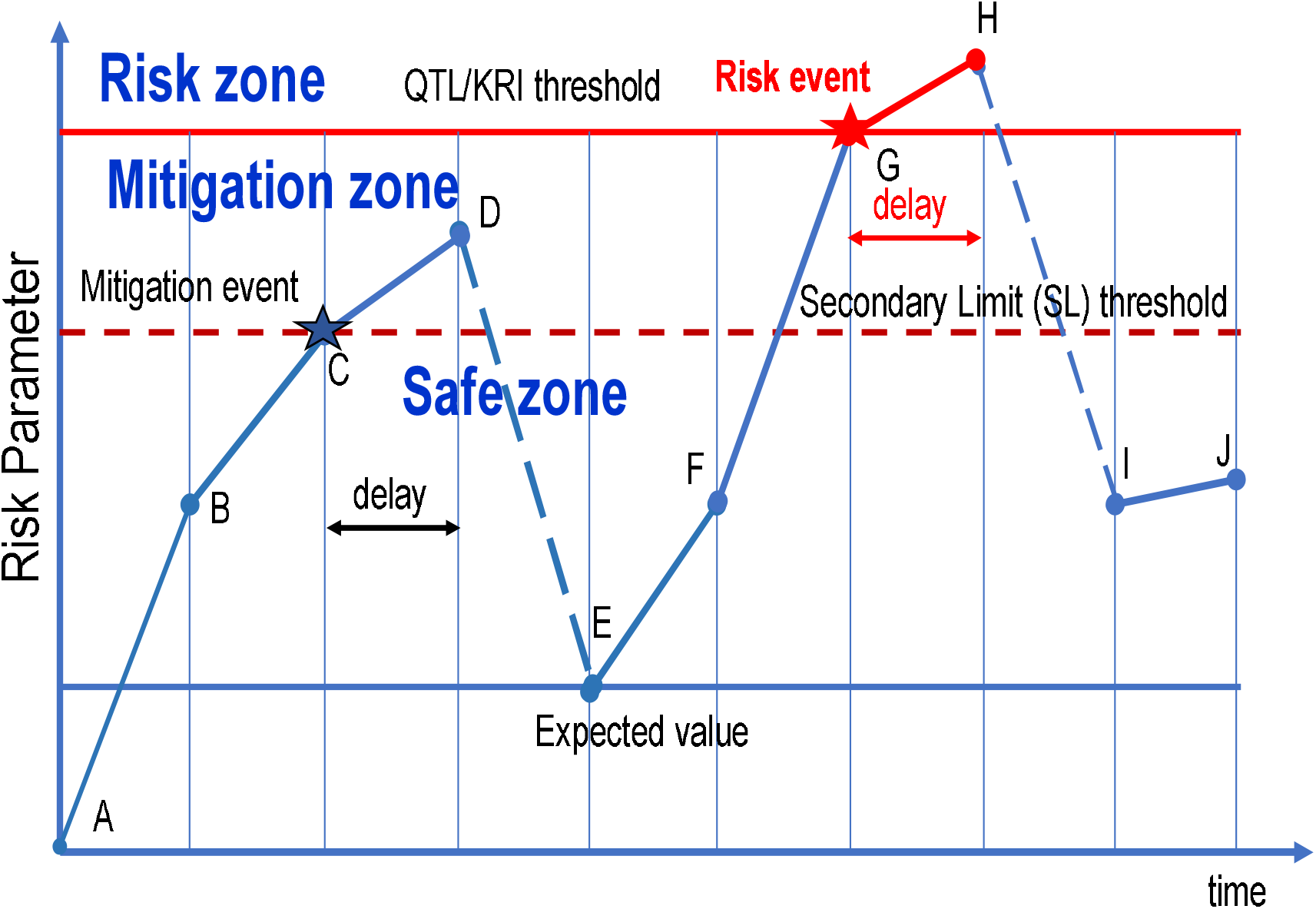
Illustration of RP dynamics. Legend: X-axis—time, Y-axis—the dynamic of an RP parameter.

The RP space includes several horizontal lines: The “expected value” line for the RP, the SL line, and the QTL line. The space between the SL and the QTL is the mitigation zone, and the space between the SL and the X-axis is a safe zone.

The RP dynamics can be described as follows. When the RP reaches or exceeds the SL line (point C – mitigation event), the mitigation should start according to the mitigation plan with a delay—the longer the delay, the closer an RP is to the QTL threshold (point D) and the early warning effect is reduced. The mitigation plan includes training, additional site visits, teleconferencing calls, opening backup sites, and other actions depending on the risk, time, and costs = **C**_**DE**_. As a result of the mitigation, the RP value is reduced to the expected value (point E), consuming contingency resources. Then, for illustration purposes, the rise of RP is very steep and may reach (or exceed) the QTL line (point G - risk event). Due to the delay, the mitigation can only start at the point H. The mitigation costs include planned **C**_**HI**_ and unplanned costs - **ΔC**_**HI**_ (please see the fuel indicator example above, wasting time and higher fuel costs). The reduction of RP may not reach the RP Expected Value level due to insufficient contingency funds. The total mitigation costs are -

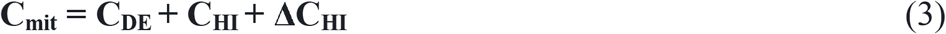

The model outcome includes two essential parameters.

1. Risk as the probability of RP reaching QTL level and described with Equations (4) and (5)

The risk - R in the Monte Carlo simulation model is calculated as:

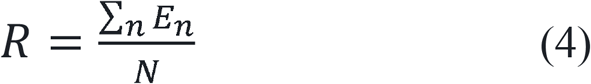

N – number of simulation cycles, n – simulation cycle where the risk event occurs. 1 ≤ n ≤ N.

*E*_*n*_ = {1- risk event occurs, i.e., *RP*_*n*_ (*t*) ≥ *QTL*; 0 - *otherwise*}

t – time, T – clinical trial duration. 0 ≤ t ≤ T.

2. Average mitigation costs

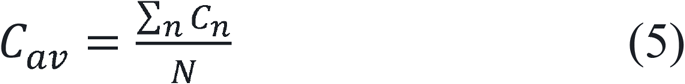

*C*_*n*_ – mitigation costs in the n-th simulation cycle.

How can QTL and SL thresholds be positioned efficiently?

#### The QTL threshold positioning

If the QTL threshold is too high, the risk event could be missed, and a “fake” risk event will be reported if it is too low. Therefore, the QTL threshold position should be aligned with the risk assessment after the mitigation. For example, if a likelihood score before the mitigation is three and after the mitigation is two, the “right” QTL position corresponds with the risk within a range (15-35%) (Table 3).

#### The SL positioning (QTL level is fixed)

Suppose the SL threshold is low (far from the QTL threshold), as presented in Figure 3A. In that case, the RP often crosses the SL threshold, triggering mitigation activities. The mitigation costs are high, but there is a low risk of reaching or exceeding QTL. If the SL threshold is high (close to the QTL threshold), the RP may cross the QTL level more often to increase the risk (Figure 3B), but mitigation costs will be low.

**Figure 3A.**
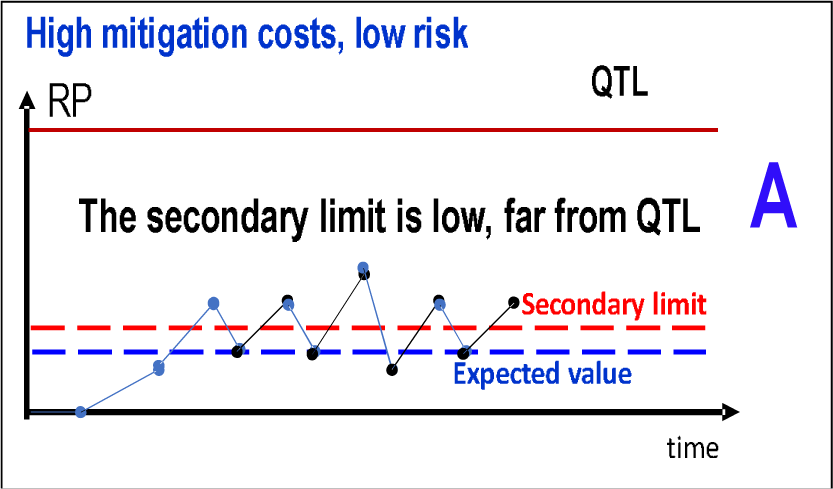
Low SL threshold.

**Figure 3B.**
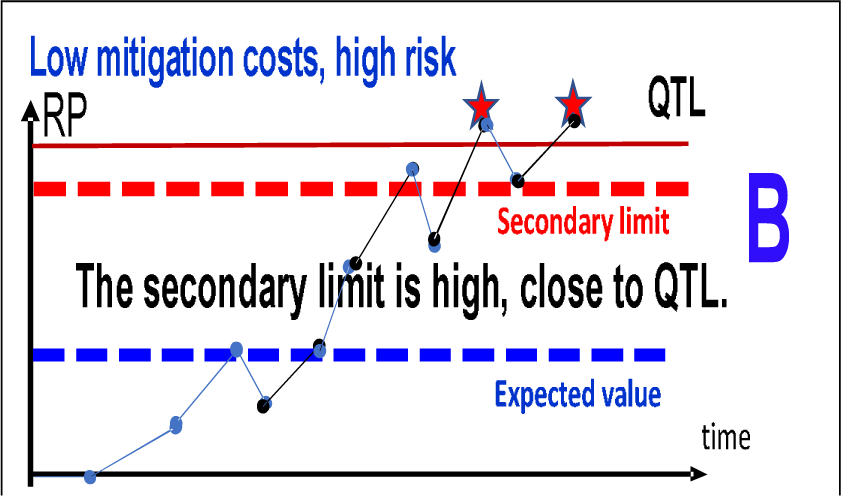
High SL threshold.

Therefore, the SL threshold position should be aligned with the mitigation costs.

Suppose the risk and mitigation costs generated by the RBM simulator are not aligned with the Risk Optimizer input data presented in Tables 2 and 3. In that case, the input data should be reviewed and corrected, and the models should be rerun with corrected data.

### Case study

#### Problem statement

- Ten significant risks were identified and assessed in a clinical trial to be mitigated.
- Each risk is characterized by its likelihood and impact.
- Each risk may have several mutually exclusive mitigation options.
- Likelihood, impact, and mitigation costs are associated with each mitigation option.
- The contingency budget is limited.

The input data (columns A-G) is presented in Table 4.

**Table 4.** Input data for the case study (data is random). Example. Risk#1 has three mutually exclusive options: (1) no mitigation, (1.1), and (1.3) with reduced likelihood. For mitigation option #1.1, mitigation costs are fifteen, and risk-adjusted mitigation costs are 3.75.

| # Risk | Likelihood,<br>L | Impact,<br>I | Total Score,<br>LxI | Mitigation<br>cost, M | Probability,<br>P | Risk-adjusted<br>Mitigation<br>costs, MxP | Selected<br>Mitigation<br>options |
| --- | --- | --- | --- | --- | --- | --- | --- |
| A | B | C | D | E | F | G | H |
| 1 | 3 | 4 | 12 | 0 | 0.5 | 0 | 0 |
| 1.1 | 2 | 4 | 8 | 15 | 0.25 | 3.75 | 1 |
| 1.2 | 2 | 4 | 8 | 45 | 0.25 | 11.25 | 0 |
| 1.3 | 1 | 4 | 4 | 120 | 0.1 | 12 | 0 |
| 2 | 4 | 4 | 16 | 0 | 0.9 | 0 | 0 |
| 2.1 | 3 | 4 | 12 | 80 | 0.5 | 40 | 0 |
| 2.2 | 2 | 4 | 8 | 105 | 0.4 | 42 | 0 |
| 2.3 | 2 | 4 | 8 | 180 | 0.25 | 45 | 1 |
| 3 | 3 | 5 | 15 | 0 | 0.5 | 0 | 0 |
| 3.1 | 3 | 5 | 15 | 60 | 0.5 | 30 | 1 |
| 3.2 | 2 | 5 | 10 | 120 | 0.4 | 48 | 0 |
| 3.3 | 1 | 5 | 5 | 165 | 0.4 | 66 | 0 |
| 4 | 3 | 4 | 12 | 0 | 0.5 | 0 | 0 |
| 4.1 | 2 | 4 | 8 | 96 | 0.3 | 28.8 | 1 |
| 5 | 5 | 4 | 20 | 0 | 0.9 | 0 | 0 |
| 5.1 | 3 | 4 | 12 | 50 | 0.5 | 25 | 1 |
| 5.2 | 2 | 4 | 8 | 132 | 0.25 | 33 | 0 |
| 5.3 | 2 | 4 | 8 | 150 | 0.25 | 37.5 | 0 |
| 5.4 | 2 | 4 | 8 | 180 | 0.25 | 45 | 0 |
| 6 | 4 | 4 | 16 | 0 | 0.75 | 0 | 0 |
| 6.1 | 2 | 4 | 8 | 69 | 0.25 | 17.25 | 1 |
| 7 | 4 | 5 | 20 | 0 | 0.75 | 0 | 0 |
| 7.1 | 1 | 5 | 5 | 90 | 0.1 | 9 | 1 |
| 8 | 4 | 4 | 16 | 0 | 0.75 | 0 | 0 |
| 8.1 | 1 | 4 | 4 | 150 | 0.1 | 15 | 1 |
| 9 | 3 | 4 | 12 | 0 | 0.5 | 0 | 0 |
| 9.1 | 1 | 4 | 4 | 135 | 0.1 | 13.5 | 1 |
| 10 | 4 | 4 | 16 | 0 | 0.5 | 0 | 0 |
| 10.1 | 2 | 4 | 8 | 150 | 0.25 | 37.5 | 1 |

### Deriving efficient risk operational rules

The goal is to efficiently position QTL and SL thresholds for each risk presented in Table 4 to be aligned with risk assessment and contingency resources.

The problem is delineated in two interconnected sub-problems.

1. Deriving an optimal risk mitigation strategy (Figure 1) [1] involves selecting the mitigation option for each risk and deriving the corresponding residual risk (Risk Optimizer). For example, for risk #1, mitigation option #1.1 was selected after the modeling. (Table 4, column H). This means that the residual risk = 2 (the probability of occurrence is within the range of 15-35%) (Table 3), and risk-adjusted mitigation costs = 3.75.
2. Deriving efficient operational rules – positioning QTL and SL thresholds for each risk aligned with risk assessment and contingency resources (RBM Simulator).

If one or both parameters (QTL and SL thresholds) are not aligned, risk assessment and mitigation resources are corrected (Figure 1, block #3). Then, the recalculation of models #1 and #2 for the adjusted thresholds (Figure 1) is necessary. Aligning QTL thresholds with risk assessment and mitigation resources may require up to four modeling cycles.

#### Efficient positioning of QTL and SL thresholds

The RBM simulator aims to validate the efficiency of selected risk mitigation strategy and derive efficient operational rules.

#### Input data

a. A range of acceptable QTL thresholds based on historical trials, experience, and expert opinion similar to [13].

**QTL**_**min**_ ≤**QTL** ≤ **QTL**_**max**_.

Some QTL thresholds are fixed, e.g., “*<u>% or number of randomized study participants who do not meet exclusion/inclusion criteria “[4]</u>*. In this case, only the SL threshold should be positioned.

b. Risk assessment range, e.g., see Table 3.
c. Risk parameter deviation (min, max)
d. Risk Parameter reduction due to the mitigation (min, max)
e. Cycle time to resolve the mitigation or risk signal (delay)
f. Cost per mitigation action (min, max)

### Modeling results

#### 1. Efficient QTL threshold positioning. Baseline scenario

Let’s consider the positioning of the QTL threshold for risk #1. Risk #1 was mitigated as part of the risk mitigation strategy for the entire portfolio of risks (Table 4) using the model presented in [1]. Mitigation option #1.1 was selected. The risk likelihood was reduced from = 3 to = 2 or, on average, from 0.5 to 0.25, with a range of 0.15-0.35 (Table 3).

Simulation experiments were conducted for the QTL threshold range (0.12 – 0.19) and SL range from 0.06 to the level of the QTL threshold. The goal is to find a combination of QTL and SL thresholds aligned with the clinical trial risk and contingency resources.

If the QTL threshold is 0.12, the risk is outside of the required range of the likelihood of 0.15-0.35 for all SLs from 0.06 to 0.12.

Increasing the QTL threshold reduces the risk. If QTL = 0.15, the risk for all SLs is within the range (0.15-0.35). The points marked X correspond to SL threshold absence (reactive risk mitigation). Results in Figure 5 indicate that without SL, the risk of reaching or exceeding the QTL threshold is higher than with SL (proactive risk mitigation).

**Figure 5.**
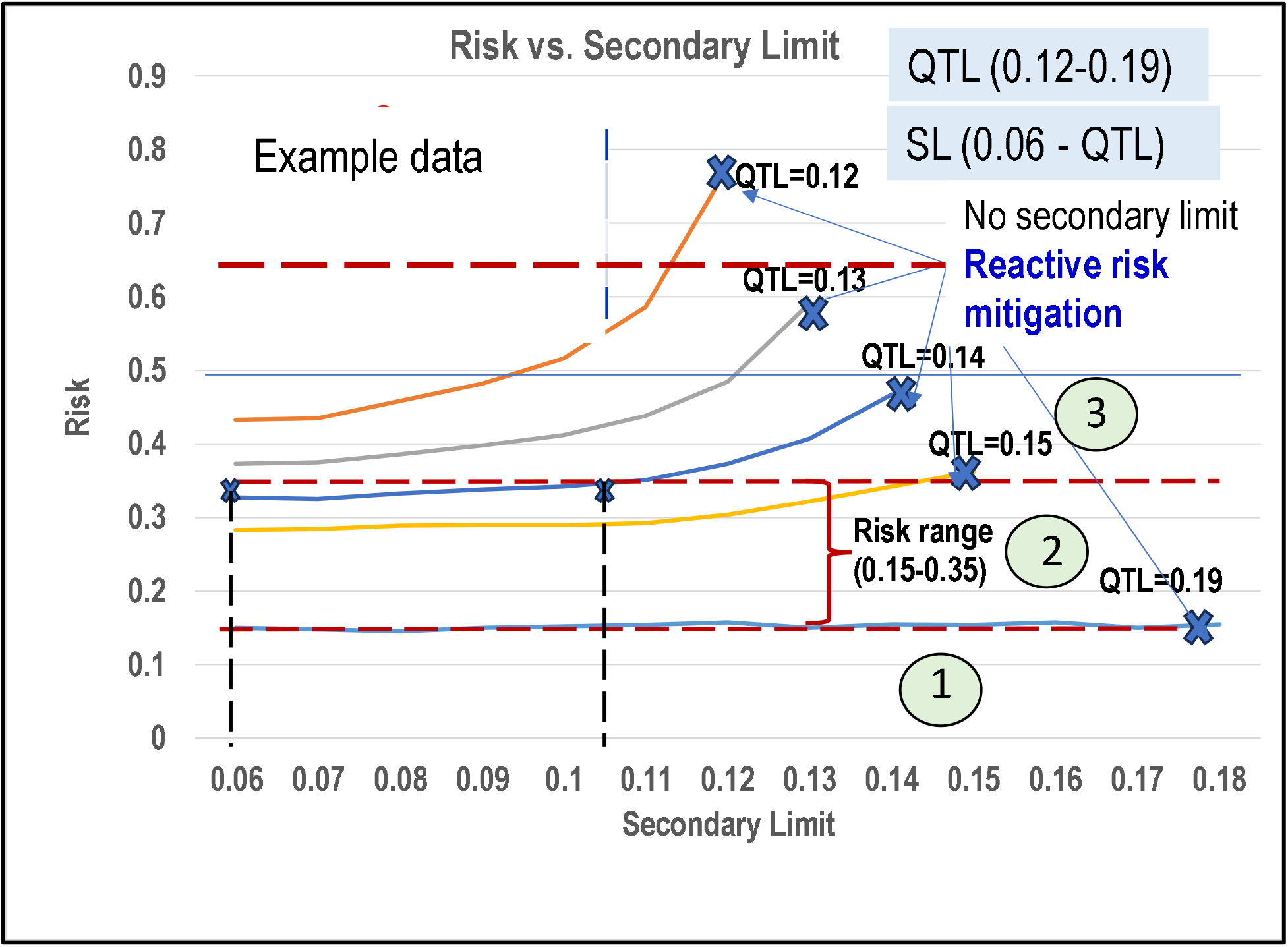
Clinical trial risk vs. SL for different QTL values. ***Legend***. *The horizontal lines are boundaries between risk zones. Example: Zone #2 corresponds to the risk range of 0*.*15 – 0*.*35* (*Table 3*).

#### 2. Efficient Secondary Limit (SL) threshold positioning

Simulation experiments were conducted for a fixed QTL threshold = 0.15 and variable SL. They are presented in Figure 6, capturing SL vs. risk-adjusted mitigation costs according to the observations in Figures 3A and 3B.

**Figure 6.**
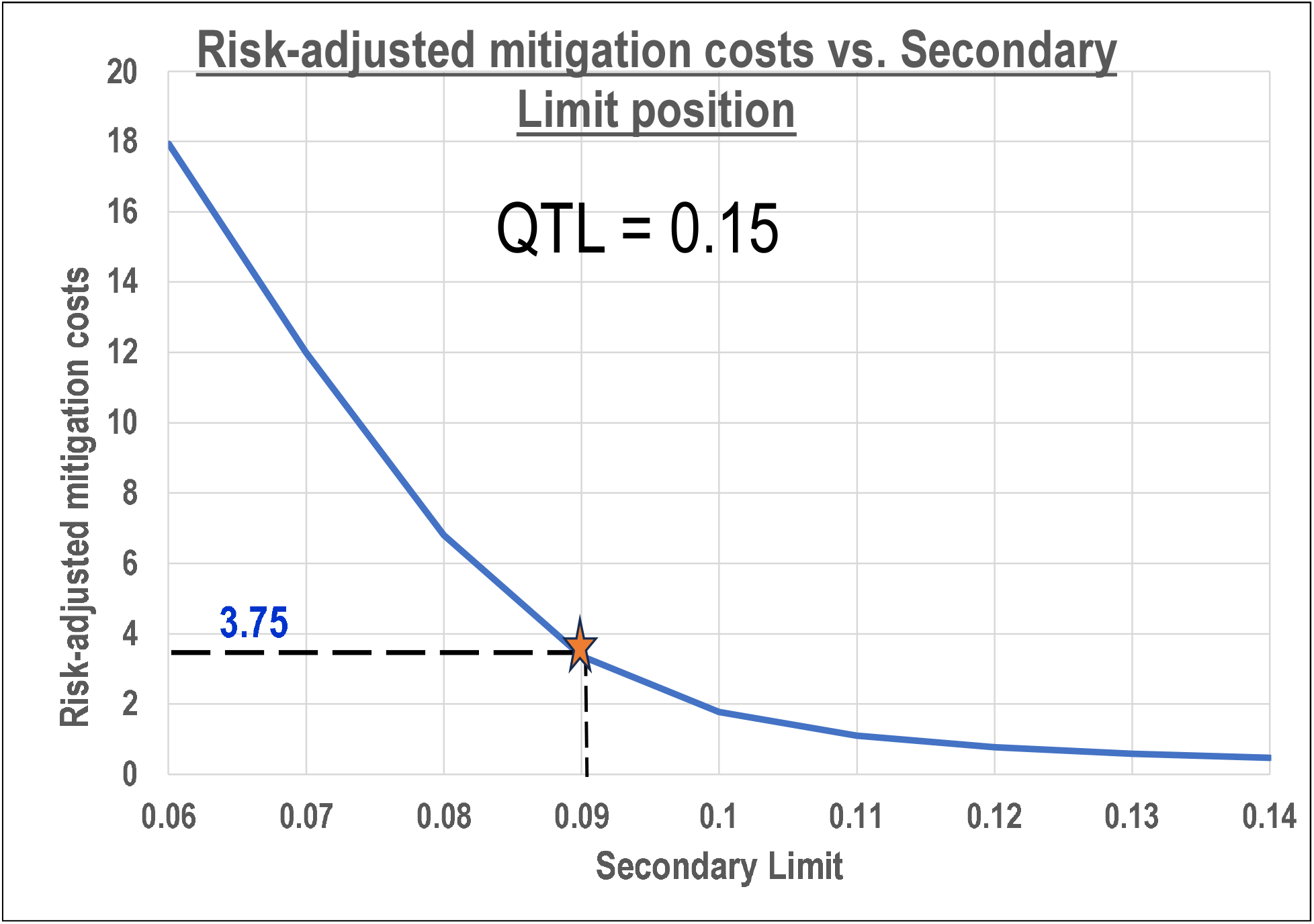
Risk-adjusted mitigation costs vs. Secondary Limit position.

The graph indicates that the lower the SL, the higher the mitigation costs. Selected risk mitigation option 1.1 for risk #1 requires a risk-adjusted mitigation cost of 3.75, corresponding to SL = 0.09, QTL = 0.15, and risk =0.29 (within the range 0.15-0.35, Figure 5).

Optimization and simulation models must be recalculated if modeling results (risk and mitigation costs) are not aligned with established QTL and SL limits (Figure 1). This process requires an expert opinion, especially in the risk reassessment.

One of the model’s critical parameters is the “cycle time response to the risk signal.” A benchmarking study [15] showed that the average time to respond to the risk signal was around 35 days. Is it too long, too short, or just right? It seems intuitive that the cycle time should be as little as possible because an RP could continue to grow after crossing the SL threshold and potentially jeopardize the integrity of trial results.

The graph in Figure 7 indicates that the longer the delay between the risk signal and the response, the higher the probability of a risk event. Without response after crossing the SL threshold, the RP will continue increasing until the start of the mitigation.

**Figure 7.**
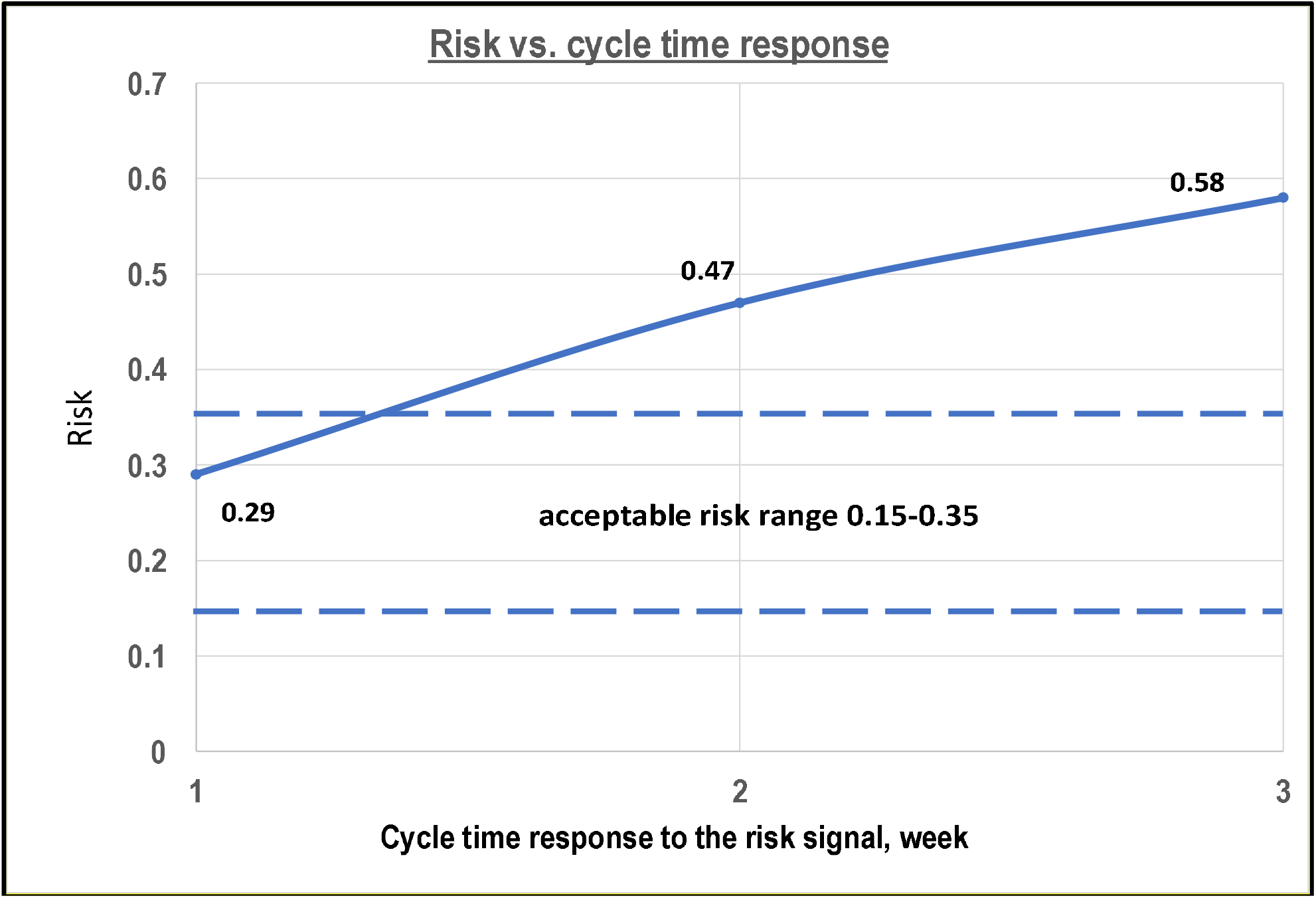
Risk vs. cycle time response to the risk signal.

According to the graph in Figure 7, the risk = 0.29 is within an acceptable range (0.15-0.35) only if the delay in response to the risk signal is about a week. If the mitigation postponement equals two weeks, the risk becomes 0.47; for a three-week delay, the risk becomes 0.58. If the delay cannot be reduced to the required value, then the only option is to raise the QTL threshold to stay within risk limits.

## Conclusion

The paper presents a technique for efficiently positioning QTL and Secondary Limit thresholds. The technique advantages:

- It is clinical trial specific.
- It is based on the combined usage of optimization and simulation models
- It aligns thresholds with risk assessment and contingency resources.
- Small and medium companies can use it without utilizing substantial historical data.
- The thresholds can be recalculated during clinical trial execution.

## Data Availability

Data is random to illustrate the technique

## Conflict of interest

No conflict of interest was reported.

The author thanks Steve Young, CTO of Clue Points, Inc., for the helpful discussions.

## Footnotes

1 A risk mitigation strategy is the coordinated selection of the most efficient mitigation action for each risk in a portfolio, aligned with risk assessment, limited contingency resources, and risk tolerance.

2 Risk operational rules are a set of algorithms that execute a risk mitigation strategy during a clinical trial.

3 [3] also stated that RP could be an absolute metric.

4 The Quality Tolerance Limit (QTL), a predefined threshold, is associated with the RP. Its breach can severely impact patients’ safety and the reliability of clinical trial results, underscoring its significance.

5 The Key Risk Indicator (KRI) is a predefined threshold associated with the RP where the integrity of a trial’s results can be impacted.

6 From the modeling point of view, there is no differentiation between QTL and KRI thresholds at the clinical trial level.

7 Since the technique for positioning QTL and KRI thresholds is the same, only the QTL term will be used further in the paper.

8 Secondary Limit (SL) – a threshold that should trigger mitigation activities to prevent the RP from reaching the QTL/KRI and avoid systemic risk.

9 Additional research is needed to benchmark both parameters robustly across diverse types of clinical trials.

